# Decoding the Genetic Architecture of Autistic Traits in the Aging Population

**DOI:** 10.64898/2026.06.10.26355340

**Authors:** Panhui Tian, Xiaoli Rao, Yang Sui, Shilin Gao, Yingying Meng, Xu Han, Tianyun Wang

## Abstract

Autism research has mostly focused on diagnostic frameworks in childhood. However, autistic traits including social skills, communication, attention switching, attention to detail, and imagination, may also vary in many undiagnosed individuals beyond childhood, and the genetic architecture of autistic traits in undiagnosed aging adults remains poorly understood. Here, we performed an exome wide association study of autistic traits in adults aged ≥40 from the UK Biobank (n = 161,269) and independently validated key findings in the SPARK cohort (n = 142,357). We identified exome wide significance at 17q21.31, represented by a lead variant associated with social skills (rs199533, β = 0.081, P = 2.04 × 10^−11^). In addition, we identified an independent signal for communication (rs12632110, β = 0.042, P = 3.07 × 10^−12^) and two independent signal for attention switching (rs690733, β = 0.046, P = 4.26 × 10^−12^; rs2164272, β = −0.047, P = 1.73 × 10^−12^). Gene-based analyses further implicate loss-of-function variation in *ZSCAN2* (β = 1.00, P = 2.44 × 10^−6^), which associated with communication differences. Enrichment analyses reveal preferential expression of implicated genes in the cerebral cortex, while phenotypic and neuroimaging analyses link those variants to cortical brain structure and regional volume. Taken together, these findings delineate the genetic architecture of autistic traits in the aging population and link genetic variation to downstream molecular and neuroanatomical mechanism.

## Introduction

Autism spectrum disorder (ASD) is defined as a neurodevelopmental condition typically diagnosed in childhood^1,2^. Yet, its core features, social communication difficulties and restricted, repetitive behaviors, are not confined to childhood, and often persist into adulthood and later life, influencing cognition, social functioning, and emotional experience across the lifespan^3,4^. While genetic research has made landmark strides in pediatric and familial ASD studies^5–8^, a critical paradox emerges as the vast majority of genetic studies are anchored to a childhood diagnosis, yet autistic traits are widely distributed in the population and extend far beyond clinically diagnosed children. This creates a profound blind spot. The very individuals who could illuminate how autistic traits manifest, the general adult and aging population, particularly those who have navigated life without a clinical diagnosis, have been systematically excluded from genetic inquiry^9–12^. We know remarkably little about the genetic underpinnings of autistic traits in this majority population. This is not a niche oversight, it is a fundamental gap in understanding the full architecture of this complex human phenotype^10^.

This knowledge gap is now a pressing demographic and scientific imperative. With increasing life expectancy and the advent of an aging society, a vast ‘invisible’ cohort is emerging that adults with subclinical autistic traits who have navigated life without a clinical diagnosis. This undiagnosed majority is now aging, yet their experiences and challenges remain profoundly unrecognized by a healthcare and research paradigm anchored to childhood presentation. For this undiagnosed majority, the genetic architecture of their traits is largely unknown. So here, in order to address this foundational deficit, we perform a large-scale exome wide association study (EWAS) of quantitatively measured autistic traits in adults with age over 40 from the general population, the vast majority of whom lack an ASD diagnosis. By integrating deep exome sequencing data from 161,269 UK Biobank participants with dimensional phenotypic data from the Autism-Spectrum Quotient (AQ)^13^, we shift the paradigm from a binary, diagnosis-bound framework to a continuous, lifespan-oriented one.

By illuminating the genetic architecture of autistic traits in the general older adult population, this work provides an essential biological framework for understanding autism as a lifelong condition. This is a critical step towards a future where understanding and supporting for autistic individuals are not confined to childhood, but extend across the entire human lifespan.

## Results

### Exome-wide association analysis for autistic traits

After stringent population-level filtering (see Methods), we retained participants who completed all 50 items of the AQ questionnaire and were ≥40 years old, yielding a final sample set of 131,005 individuals. Then, we conducted exome-wide association studies to assess associations between variants and the AQ total score as well as its five subdomains. In addition, MAGMA gene-based analyses were performed for common variants to evaluate gene-level associations.

In the common-variant EWAS, we identified 65 SNPs reaching genome-wide significance across three AQ subdomains: social skill, communication, and attention switching. Among these, 64 SNPs represented novel associations, while only one SNP (rs12373168) had been previously reported. Notably, 59 of the 65 genome-wide significant SNPs were associated with social skill and mapped to a well-established locus at chromosome 17q21.31, which harbors a common inversion polymorphism and has been repeatedly implicated in neurodevelopmental and neuropsychiatric traits^14–17^. These SNPs formed a large continuous linkage disequilibrium (LD) block, as confirmed by regional association plots by using LocusZoom (Fig. S5). LD clumping and conditional analyses indicated that these variants represent a single association signal, with rs199533 as the lead SNP.

In addition, two SNPs were associated with communication, rs1046956 (β = 0.0423, P = 3.25 × 10⁻□) and rs12632110 (β = 0.0423, P = 3.07 × 10⁻□), which were both mapped to *SEMA3F*. And four SNPs were associated with attention switching, among which three of these variants, rs3808513 (β = −0.0461, P = 3.36 × 10⁻□), rs2164272 (β = −0.0471, P = 1.73 × 10⁻□), and rs2293855 (β = −0.0460, P = 3.72 × 10⁻□) were in strong LD and localized to *MTMR9*. As for the remaining variant, rs690733 (β = 0.0455, P = 4.26 × 10⁻□) was located within *EXD1*.

We further conducted gene-based analyses using MAGMA on summary statistics derived from the common-variant EWAS. A total of 23 genes were identified as significantly associated with four autistic trait domains (Fig. S6, Table S6). Notably, *SEMA3F* exhibited robust gene-level associations not only with communication, but also with social skill and the overall AQ score (Fig. S11). This cross-domain convergence highlights *SEMA3F* as a potential pleiotropic contributor to multiple dimensions of autistic traits in adulthood.

Rare variants were categorized into loss-of-function (LoF) variants and missense variants with MPC score ≥ 2, and were analyzed in relation to the AQ total score and its five subdomains. As an initial step, we assessed the overall burden of different rare variant classes, including LoF, MPC ≥ 2, LoF + MPC ≥ 2, and synonymous variants, across all six traits. We found that both LoF variants and LoF + MPC ≥ 2 variants were significantly associated with increased scores in attention switching, attention to detail, imagination, and the AQ total score, indicating a substantial contribution of rare functional variants to autistic traits (Fig. S2).

We then conducted gene-based association analyses using SKATO test. LoF variants in *NBR1* were significantly associated with increased AQ total score (β = 1.76, P = 1.91 × 10⁻□), while LoF variants in *ZSCAN2* were associated with increased communication scores (β = 1.00, P = 2.44 × 10⁻□).

### Interaction analysis, leave-one-variant-out analysis and conditional analysis

To assess potential effect modification by sex and age, we conducted interaction analyses for all 65 common variants identified in the exome wide association analyses. For each variant, multiplicative interaction terms with sex and age were tested. None of the common-variant associations showed statistically significant SNP×sex or SNP×age interaction effects after correction for multiple testing (Tables S4-S5), indicating that the detected common-variant effects on AQ traits were largely consistent across sex and age strata.

We next evaluated interaction effects for the two rare-variant gene-based associations identified in the primary analysis. For LoF variants in *NBR1*, which were associated with AQ total score, no significant gene×sex interaction was observed (P = 0.93). However, a nominally significant gene×age interaction was detected (P = 0.026), suggesting age-dependent modulation of the *NBR1* LoF effect on AQ total score. Similarly, for LoF variants in *ZSCAN2*, which associated with communication, no significant interaction with sex was observed (P = 0.51), whereas a nominal gene×age interaction was identified (P = 0.033) (Table S7). These results indicate that rare-variant associations were not sex-specific but may vary with age.

To assess whether the rare-variant gene-based signals were attributable to individual LoF variants, we performed LOVO analyses for *ZSCAN2* and *NBR1* (Fig. S10). For *ZSCAN2* (LoF), sequential removal of contributing variants resulted in modest changes in the gene-level evidence (|Δ−log_10_(P)| < ∼2), with the largest attenuation observed upon excluding 15:84621601:CAG:C (|Δ−log_10_(P)| = -1.79), indicating that the association is distributed across multiple variants rather than dominated by a single variants (Fig. S10a).

In contrast, the *NBR1* (LoF) association showed marked dependence on a single variant. Exclusion of 17:43200252:GGAGGAGGAT:G resulted in a substantial attenuation of the gene-level association (|Δ−log_10_(P)| = −5.41), whereas removal of other variants produced only minor changes (|Δ−log_10_(P)| ≈ 0) (Fig. S10b). This pattern indicates heterogeneity in variant-level contributions and suggests that the observed *NBR1* LoF association is largely driven by one influential LoF variant within the set.

To further resolve the independence of association signals within each autistic trait, we performed LD clumping followed by conditional analyses for all genome wide significant regions. For the social skill domain, although 59 SNPs reached genome-wide significance at the 17q21.31 locus, LD clumping identified only one single lead SNP (rs199533). Conditional analysis adjusting for rs199533 abolished all genome-wide significant associations within the region, indicating that the observed signal is driven by a single underlying association (Fig. S13). For the communication domain, two genome-wide significant SNPs were identified. LD clumping indicated that rs12632110 represents the lead signal. Conditional analysis demonstrated that no additional variants remained genome-wide significant after adjusting for this SNP, suggesting a single independent association at this locus. (Fig. S14) For attention switching, four genome-wide significant SNPs were identified. LD clumping resolved these into two independent lead SNPs, rs690733 and rs2164272. Notably, rs2164272 was associated with decreased attention switching scores, suggesting a potential protective effect. Conditional analyses further confirmed that no additional genome-wide significant variants remained after accounting for these lead SNPs, supporting the presence of two independent association signals with attention switching (Fig. S15).

Finally, to evaluate whether the rare-variant associations could be explained by nearby common variation, we conducted regional conditional analyses. Common-variant association scans were performed within ±500 kb of each implicated gene, followed by LD clumping to identify independent variants (P < 1×10⁻□, r² < 0.01). No common variants met the clumping criteria in the regions surrounding *NBR1* (AQ total score; minimum regional P = 2.97×10⁻³) or *ZSCAN2* (communication; minimum regional P = 8.28×10⁻³). Consequently, no independent common variants were available for inclusion as conditioning covariates in the rare-variant gene-based models, supporting the interpretation that the observed rare-variant associations were not attributable to local common-variant effects.

### Cross cohort validation in the SPARK cohort

To evaluate the generalizability of genetic associations identified in the UK Biobank, we performed independent validation in the SPARK cohort. Given differences in behavioral instruments between cohorts, we adopted a domain-matched strategy, aligning AQ subdomains with conceptually related SPARK measures. Specifically, variants associated with AQ social skills and communication were evaluated using the Social Communication Questionnaire (SCQ), whereas variants associated with attention-related domains were assessed using the Repetitive Behavior Scale-Revised (RBS-R) questionnaire.

We first examined replication of the four independently associated signals identified in the UK Biobank (social skills: rs199533; communication: rs12632110; attention switching: rs2164272 and rs690733). For attention switching, the two independent lead SNPs (rs2164272 and rs690733) were successfully replicated in SPARK. These variants exhibited opposite directions of effect on RBSR scores, consistent with their respective associations in the UK Biobank. Notably, rs2164272 was associated with reduced trait scores, suggesting a potential protective effect (Fig. 4b, 4d). As RBSR measures were available only in diagnosed individuals, replication for attention switching was restricted to the affected subgroup.

**Figure 1.**
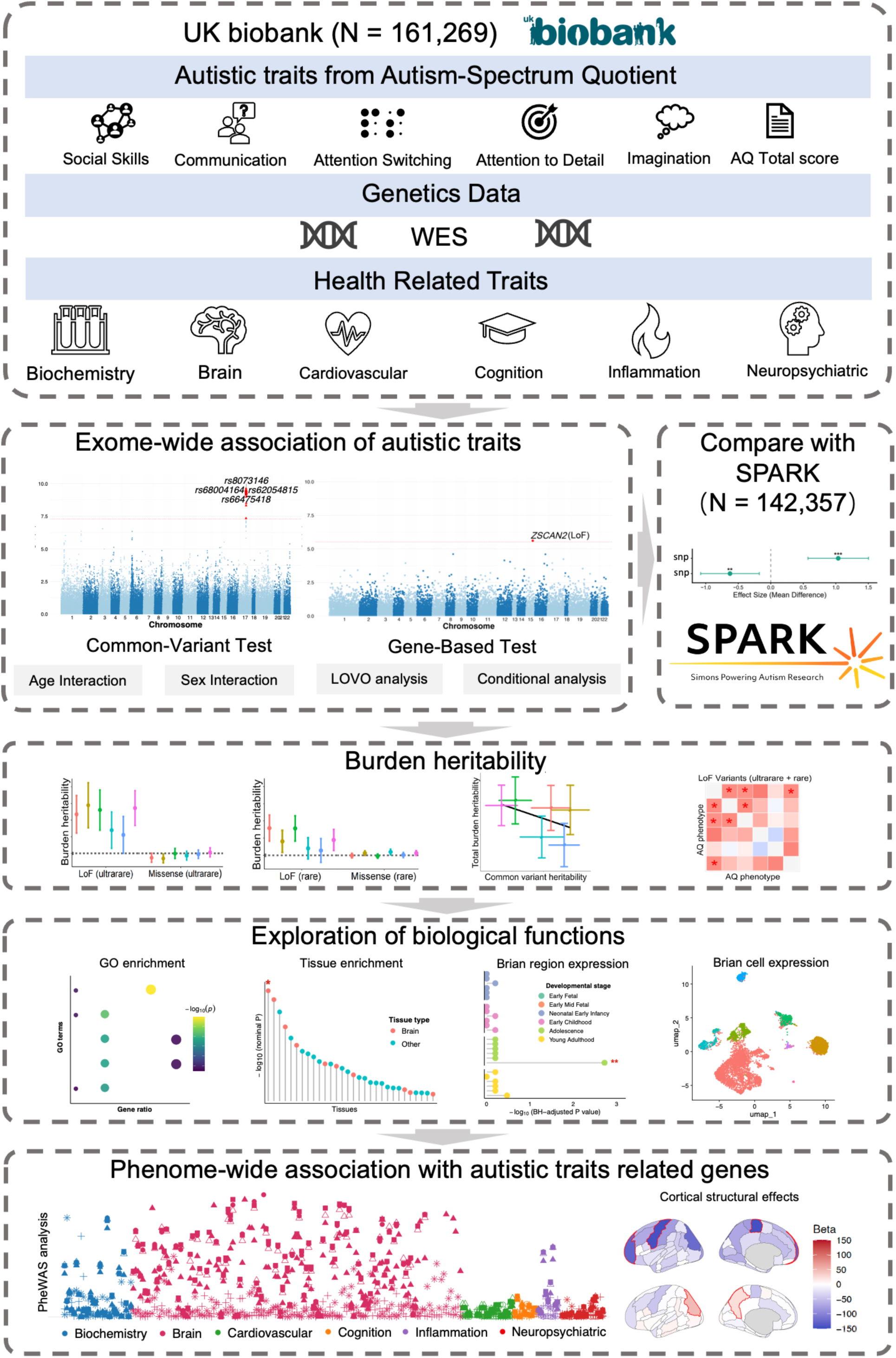
Study workflow. Research pipeline integrating data from UK Biobank (N = 161,269) and SPARK (N = 142,357) cohorts. First we extract six autistic trait dimensions from Autism Spectrum Quotient: Social Skills, Communication, Attention Switching, Attention to Detail, Imagination, and AQ Total score. These traits are combined with whole-exome sequencing and health-related data. Analytic steps include: exome-wide association studies; cohort comparison; common and gene-based variant testing; age/sex interaction analyses; leave-one-variant-out analysis; burden heritability estimation; and biological exploration via GO enrichment, tissue expression, and brain cell-type analysis. Finally, phenome-wide associations with health traits are examined.

**Figure 2.**
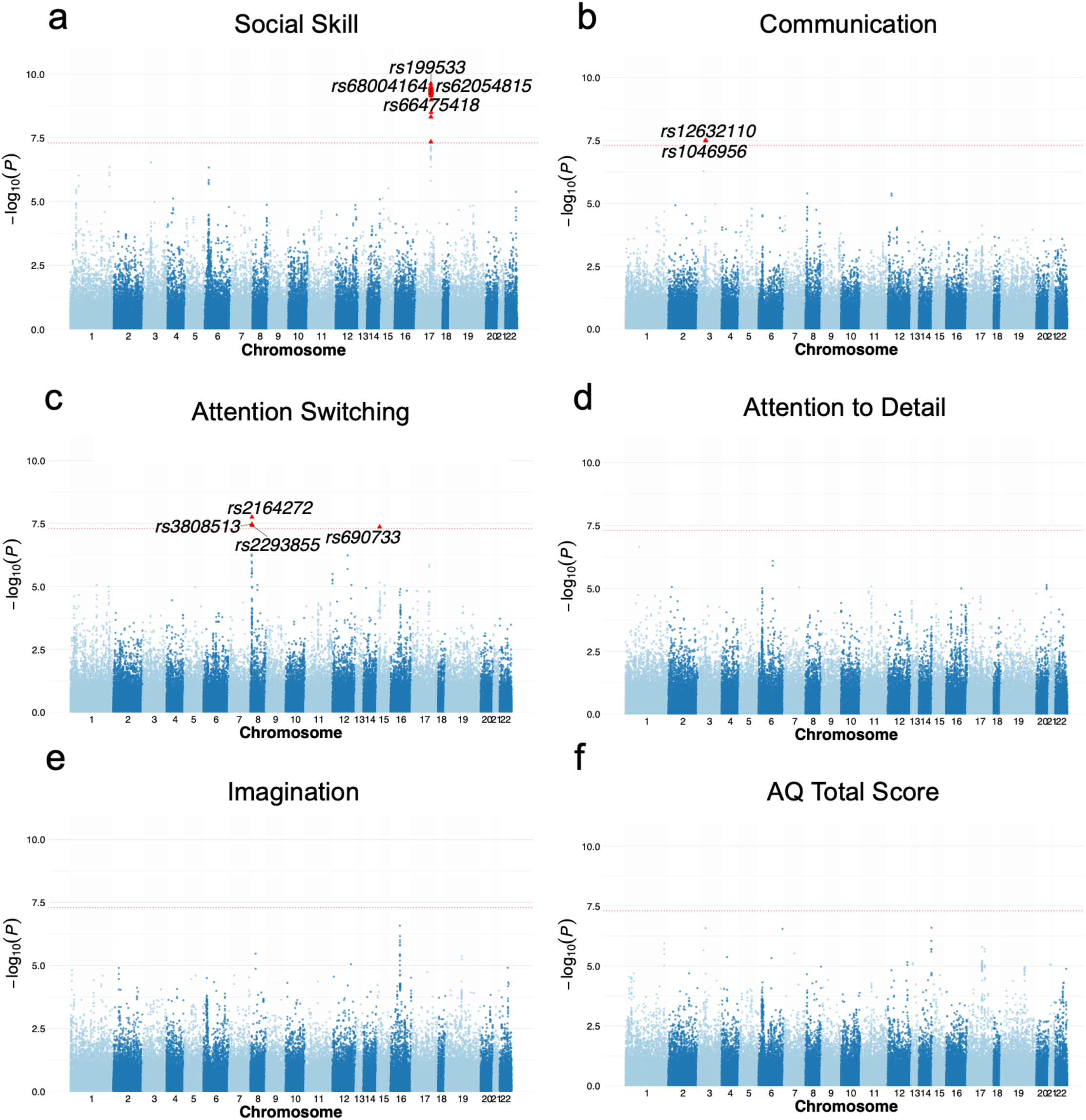
Exome-wide association analyses of common variants with autism spectrum quotient total score and subdomains. Manhattan plots showing exome-wide association results for common variants with the AQ total score and its five subdomains: Social Skill, Communication, Attention Switching, Attention to Detail, and Imagination. Each point represents a common SNV located within protein-coding regions, plotted according to its chromosomal position. For visualization clarity, only the five most significant variants in each analysis are highlighted and annotated with their rs identifiers.

**Figure 3.**
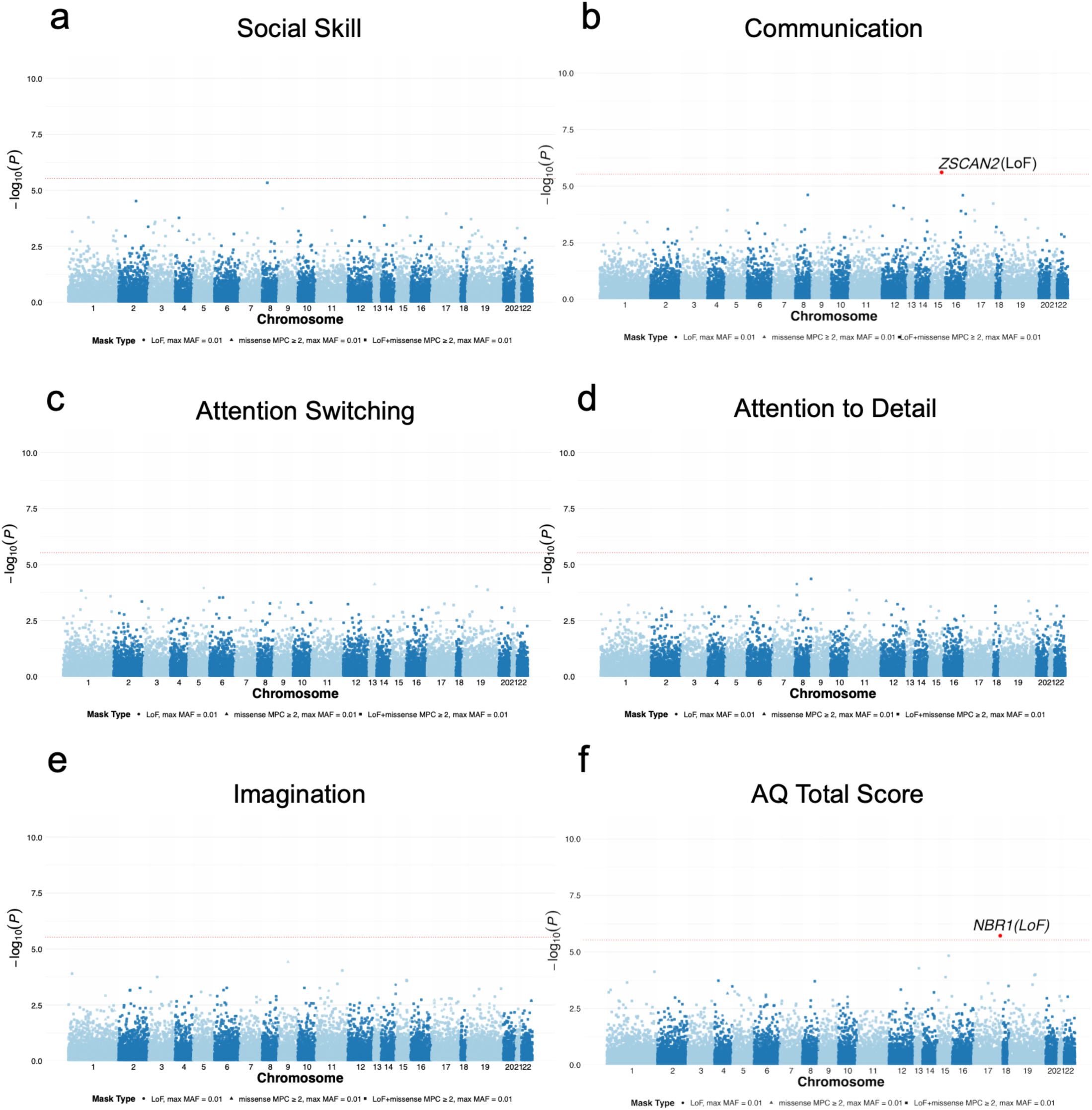
Exome-wide rare-variant gene-based association analyses of autism spectrum quotient traits. Manhattan plots showing results from exome-wide gene-based association analyses of rare coding variants with AQ subdomains and total score, including Social Skill (a), Communication (b), Attention Switching (c), Attention to Detail (d), Imagination (e), and AQ Total Score (f). Gene-based association tests were performed using SKAT-O, aggregating rare variants within protein-coding genes. Each point represents a gene, plotted according to its chromosomal position. The horizontal red dashed line indicates the Bonferroni-corrected significance threshold accounting for the total number of genes tested.

**Figure 4.**
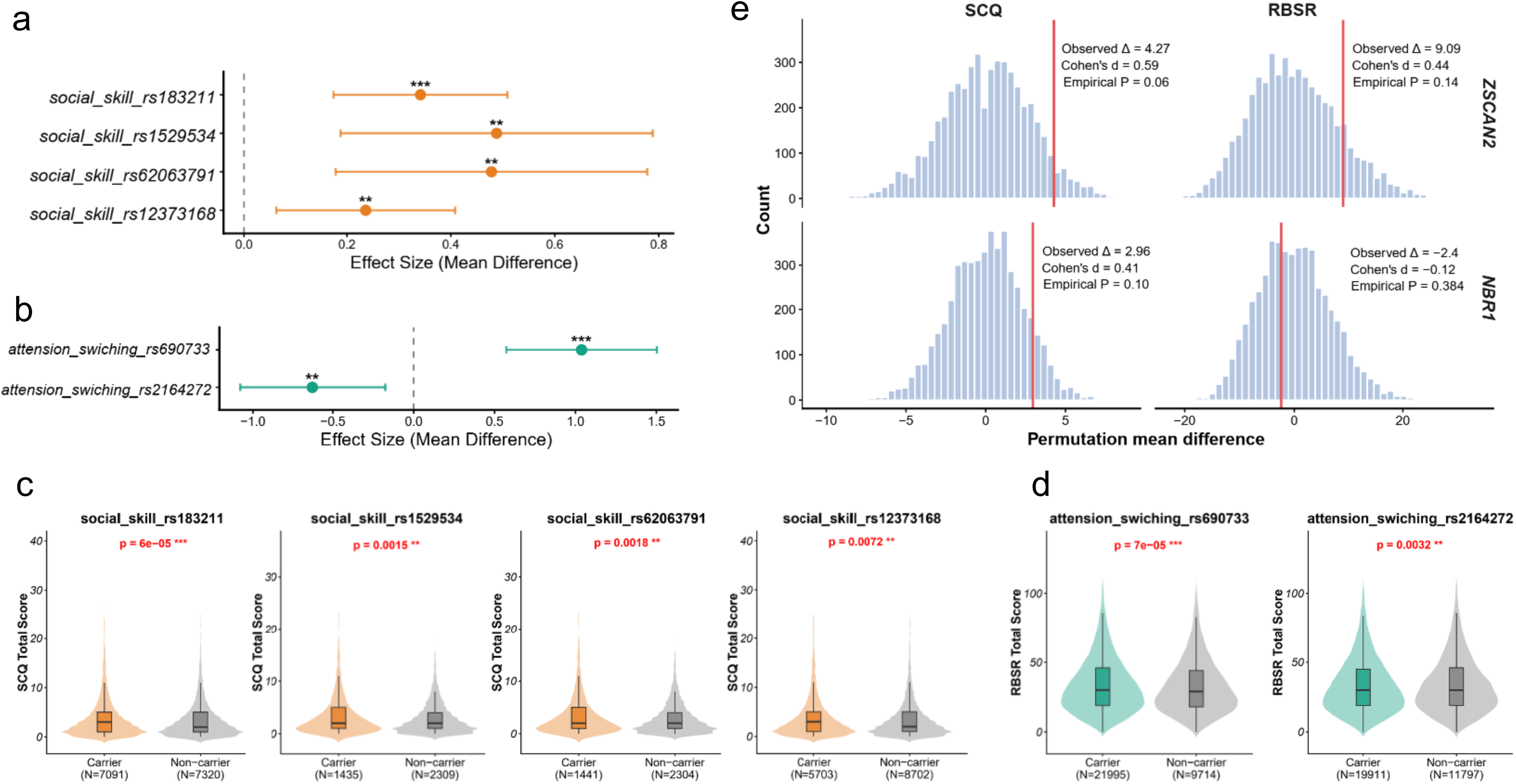
Cross cohort validation of UK Biobank genetic associations in SPARK. Common and rare genetic associations identified in the UKB using the Autism Spectrum Quotient were evaluated in the SPARK cohort using independent phenotype measures. (a, b) Effect sizes (mean differences between carriers and non-carriers) for selected common variants identified in the UKB AQ analyses and tested in SPARK. Panel a shows variants associated with AQ subdomains related to social skill and communication, validated using SCQ total scores. Panel b shows variants associated with AQ attention-related subdomains, validated using RBSR total scores. Error bars indicate 95% confidence intervals. Dashed vertical lines denote no effect. (c, d) Raw phenotype score distributions corresponding to the significant associations shown in panels A and B. Violin and box plots illustrate SCQ or RBSR total scores in carriers and non-carriers, demonstrating the underlying phenotype shifts that give rise to the observed effect sizes. (e) Validation of rare loss-of-function variants in *ZSCAN2* and *NBR1* using permutation-based burden tests in SPARK. Histograms show null distributions of mean phenotype differences generated by phenotype label permutation, with observed mean differences indicated by vertical red lines. Due to the limited number of rare variant carriers, empirical P values were estimated from permutation tests. Effect sizes are reported as both raw mean differences (Δ) and Cohen’s d.

In contrast, the lead SNPs for social skills and communication did not reach statistical significance in SPARK (Table S11). Given differences in cohort composition and phenotype measurement between the two cohorts, we further evaluated replication at the locus level. For the communication domain, no consistent association signals were observed at the locus level. However, for the social skill domain, multiple variants within the same associated locus (e.g., rs183211, rs1529534, rs60263791, rs12373168) demonstrated consistent effect directions when assessed using SCQ total scores, supporting replication at the locus level (Fig. 4a). Distributional analyses further confirmed a shift toward higher SCQ scores among carriers compared to non-carriers (Fig. 4c).

Most importantly, consistent with the design of the discovery analysis in the UK Biobank, replication analyses in the SPARK cohort were conducted in stratified subgroups. Associations for social skills were observed in the non-diagnosed subgroup, with multiple variants showing statistically significant effects, whereas no comparable associations were detected in the diagnosed subgroup.

In addition to common variants, we evaluated rare loss of function variants in two genes (*ZSCAN2* and *NBR1*) that were implicated in the UK Biobank AQ rare variant gene-based analysis. Permutation based burden tests in SPARK revealed a nominal increase in SCQ scores among carriers of rare variants in *ZSCAN2* (observed Δ = 4.27, Cohen’s d = 0.59, empirical P = 0.06), and a similar trend for RBSR scores (observed Δ = 9.09, Cohen’s d = 0.44, empirical P = 0.14) (Fig. 4e). Although these associations did not reach statistical significance, the consistent direction of effect and moderate effect sizes provide supportive evidence for the involvement of *ZSCAN2* in autistic traits. In contrast, rare variants in *NBR1* did not show significant effects on either SCQ or RBSR in SPARK (SCQ: Δ = 2.96, d = 0.41, P = 0.10; RBSR: Δ = 2.24, d = 0.12, P = 0.384) (Fig. 4e). Together, these analyses provide partial independent support for both common and rare genetic associations initially identified in UK Biobank.

### Burden heritability of autistic traits

We next quantified the contribution of rare coding variation to inter-individual differences in autistic traits using burden heritability analyses. We focused on LoF variants and deleterious missense variants (MPC ≥ 2), stratified by allele frequency into ultrarare (maf < 0.001) and rare (maf < 0.01) classes, and examined their effects on the AQ total score and five subdomains.

Across all AQ traits, ultrarare LoF variants showed the largest and most consistent contributions to burden heritability (Fig. 5a). The AQ total score exhibited a substantial enrichment of heritability explained by ultrarare LoF variants, with comparable or larger effects observed for core social–communicative domains, including social skill and communication. In contrast, attentional subdomains, particularly attention to detail, showed more modest LoF-associated heritability. Rare LoF variants demonstrated a similar but attenuated pattern, indicating a frequency-dependent decay of effect size. By comparison, missense variants with MPC ≥ 2 contributed markedly less to burden heritability across all traits and frequency classes (Fig. 5a). Even in the ultrarare category, missense burden effects were weak and generally centered near 0, suggesting that disruptive coding variation rather than predicted deleterious missense changes primarily drives rare-variant genetic liability for autistic traits.

**Figure 5.**
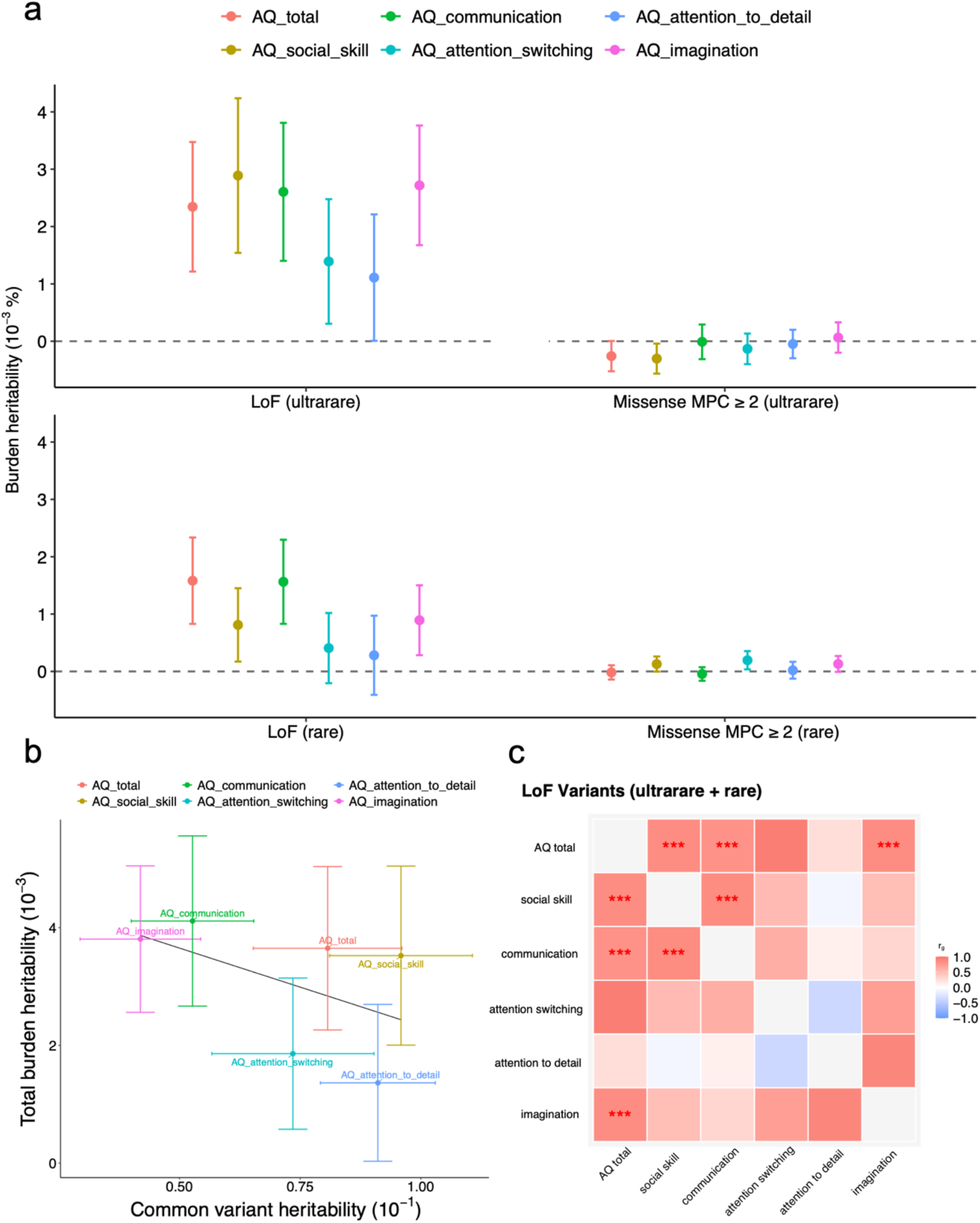
Contribution of rare coding variants to autism related traits and their relationship with common variant heritability. a, Burden heritability estimates for loss-of-function and missense MPC ≥ 2 variants across different allele-frequency classes (ultrarare and rare) for the AQ total score and five subdomains. Points represent heritability estimates and error bars denote standard errors. LoF variants show consistently larger contributions to multiple AQ traits compared with missense MPC ≥ 2 variants, particularly in the ultrarare frequency class. b, Relationship between heritability explained by common variants and total burden heritability explained by LoF variants across AQ subdomains. c, Heatmap summarizing the standardized effect sizes of LoF variant burden (ultrarare and rare combined) across AQ traits. Asterisks denote nominal significance levels (PL<L0.05, *PL<L0.01, **PL<L0.001).

To assess the relationship between rare-variant burden and polygenic background, we compared LoF burden heritability with heritability attributable to common variants across AQ subdomains. We observed an inverse relationship between these two components (Fig. 5b), whereby traits with higher common variant heritability—most notably attention to detail—showed reduced contribution from rare LoF variants. Conversely, social and communication-related traits exhibited relatively stronger rare LoF burden alongside lower common variant heritability, suggesting partially distinct genetic architectures underlying different dimensions of autistic traits.

Finally, we summarized the combined effects of ultrarare and rare LoF variants across AQ traits using standardized effect sizes (Fig. 5c). Consistent with the burden heritability estimates, social skill, communication and imagination displayed the strongest positive associations, whereas attentional traits showed weaker or more heterogeneous patterns.

### Biological functions of autistic traits related genes

To explore the potential biological mechanisms underlying our exome-wide association results, we integrated five genes harboring SPARK-replicated common variants *(MAPT, MAPT-AS1, NSF, EXD1, MTMR9)*, one identified by rare-variant gene-based analysis (*ZSCAN2*), and 23 genes prioritized by MAGMA analysis *(SEMA3F, MAS1, ITPKB, C17orf47, RAD51C, CRHR1, MAPT, STH, KANSL1, SPPL2C, NSF, PLEKHM1, ARHGAP27, ZSCAN31, MTMR9, YY1AP1, ERI1, CDK17, MSTO1, POM121L2, LRPPRC, IL27, SULT1A2),* and constructed an autistic traits associated gene set for downstream functional analyses.

Gene Ontology (GO) over-representation analysis revealed significant enrichment in biological processes related to mitochondrial localization and distribution, microtubule-mediated mitochondrial transport, and inositol phosphate metabolism (Fig. 6a). At the cellular component level, enriched terms were primarily associated with lysosomal/lytic vacuole membranes and the axolemma. At the molecular function level, significant enrichment was observed for DNA secondary structure binding and single-stranded DNA binding. Together, these results suggest that the identified genes may contribute to autistic traits through coordinated regulation of organelle positioning, membrane-associated structures, and transcriptional and protein–protein interaction processes.

**Figure 6.**
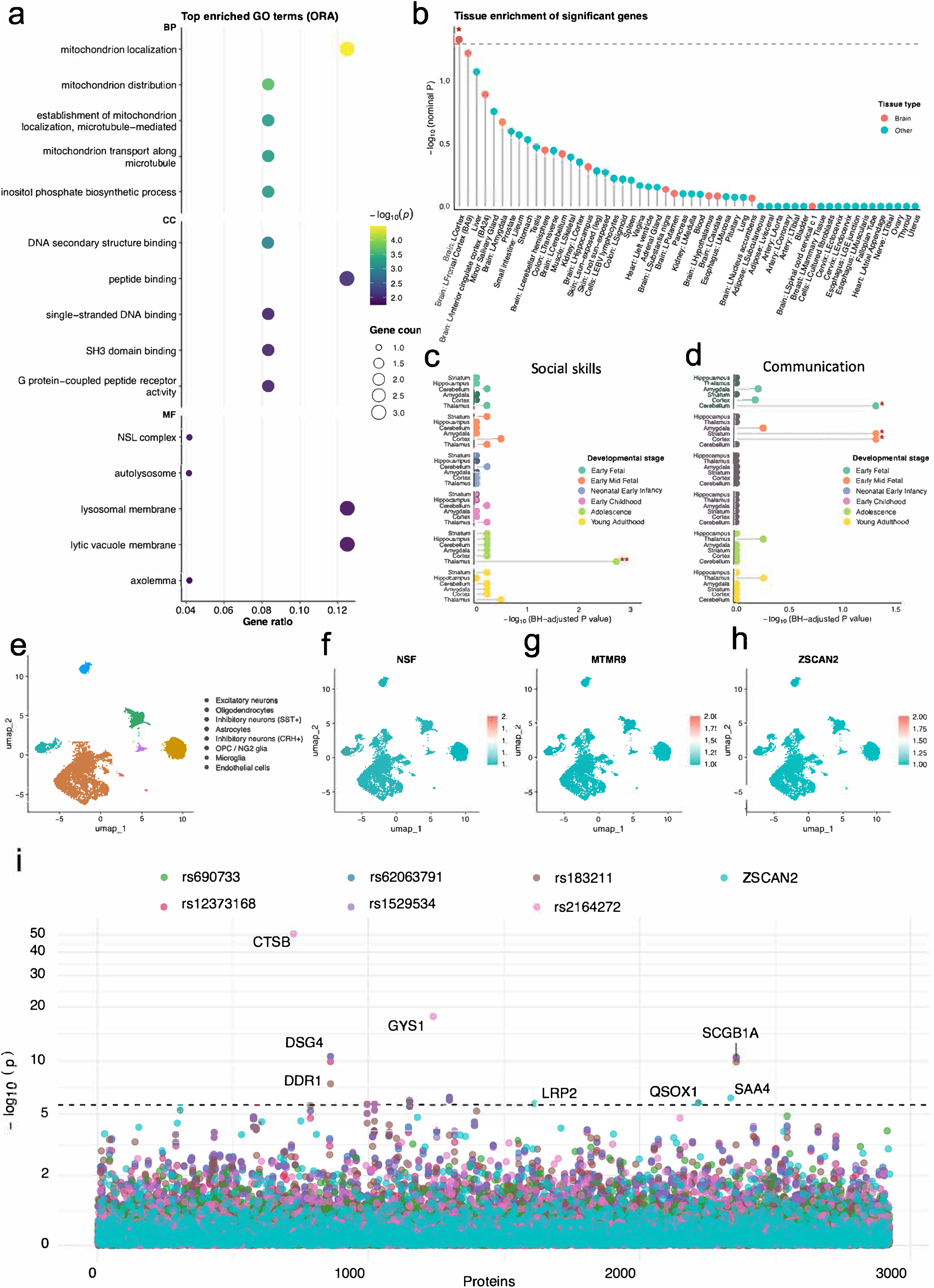
Biological functions analysis of autistic traits associated genes. a, GO analysis of genes significantly associated with social skills. b, Tissue-specific expression enrichment of significant genes based on bulk transcriptomic profiles. c, Developmental-stage-specific expression enrichment of social skill–associated genes (c) and communication-related genes (d) across human brain development using the BrainSpan transcriptome dataset. e, UMAP visualization of single-nucleus RNA sequencing data from the human cortex, with cells colored by major cell types. f–h, UMAP projections colored by normalized expression of *NSF*, *MTMR9*, and *ZSCAN2*, illustrating cell-type-specific expression patterns of representative autistic traits associated genes. i, Proteome-wide association landscape showing associations between proteins and SPARK-replicated variants and gene.

We next performed tissue-specific expression enrichment analysis using GTEx and observed that autistic traits associated genes were predominantly enriched in the cerebral cortex (Fig. 6b). Given that social impairment represents a core feature of autism, we further investigated genes associated with social skills and communication phenotypes using BrainSpan developmental transcriptomic data. Genes associated with social skills were mainly enriched in the thalamus during adolescence, whereas genes associated with communication were preferentially enriched in the cerebellum, striatum, and cortex during early fetal and early mid-fetal stages (Fig. 6c, d).

To further examine the cell-type specificity of these genes in the adult cortex, we analyzed their expression patterns in single-cell transcriptomic data from the medial prefrontal cortex (mPFC), a brain region closely implicated in social cognition. *NSF* and *MTMR9* were predominantly expressed in neurons. *ZSCAN2* showed relatively low expression in adult mPFC but was also enriched in neurons. *MAPT* and *MAPT-AS1* were broadly expressed across mPFC cell types, although *MAPT-AS1* exhibited lower expression levels. In contrast, *EXD1* showed minimal expression in adult mPFC (Fig. 6e-h, S16-S18).

To further characterize the downstream molecular consequences, we performed protein-wide association analyses using circulating proteomic data comprising approximately 2,900 proteins from the UK Biobank (Fig. 6i). This analysis identified multiple significant associations between genetic variants and circulating protein levels, with several signals surpassing stringent multiple-testing correction thresholds. Notably, associated proteins included CTSB, DSG4, GYS1, SCGB1A1, LRP2, QSOX1, and SAA4. These proteins are involved in lysosomal proteolysis, cell adhesion and membrane organization, energy metabolism, and secretory pathways, consistent with the functional pathways highlighted by genetic and transcriptomic analyses.

Together, these findings provide functional support linking genetic variation to molecular phenotypes relevant to autistic traits.

### Phenotypic association with autistic traits related genes

To further characterize the phenotypic consequences of autistic traits–associated variants and genes, particularly those supported by the SPARK cohort, we performed phenome-wide association analyses across six major phenotype domains, including neuropsychiatric, brain structural, inflammatory, cognitive, cardiovascular, and biochemical traits.

We observed that variants associated with autistic traits were predominantly enriched for brain-related phenotypes (Fig. 7a). Notably, five variants remained significantly associated with brain structural traits after Bonferroni correction, including four variants associated with social skills (rs62063791, rs183211, rs12373168, rs1529534) and one variant associated with attention switching (rs2164272). To investigate the neuroanatomical correlates of these associations, we next analyzed structural MRI data from the UK Biobank (Fig. 7b). Variants associated with social skills (rs62063791, rs183211, rs12373168, rs1529534) showed significant effects on cortical regions within the prefrontal cortex and the precentral gyrus, with risk allele carriers exhibiting reduced regional volumes in these areas. In contrast, the attention switching associated variant rs2164272 (β = −0.047, P = 1.73 × 10⁻□) was associated with increased volume of the superior division of the lateral occipital cortex, consistent with a protective effect.

**Figure 7.**
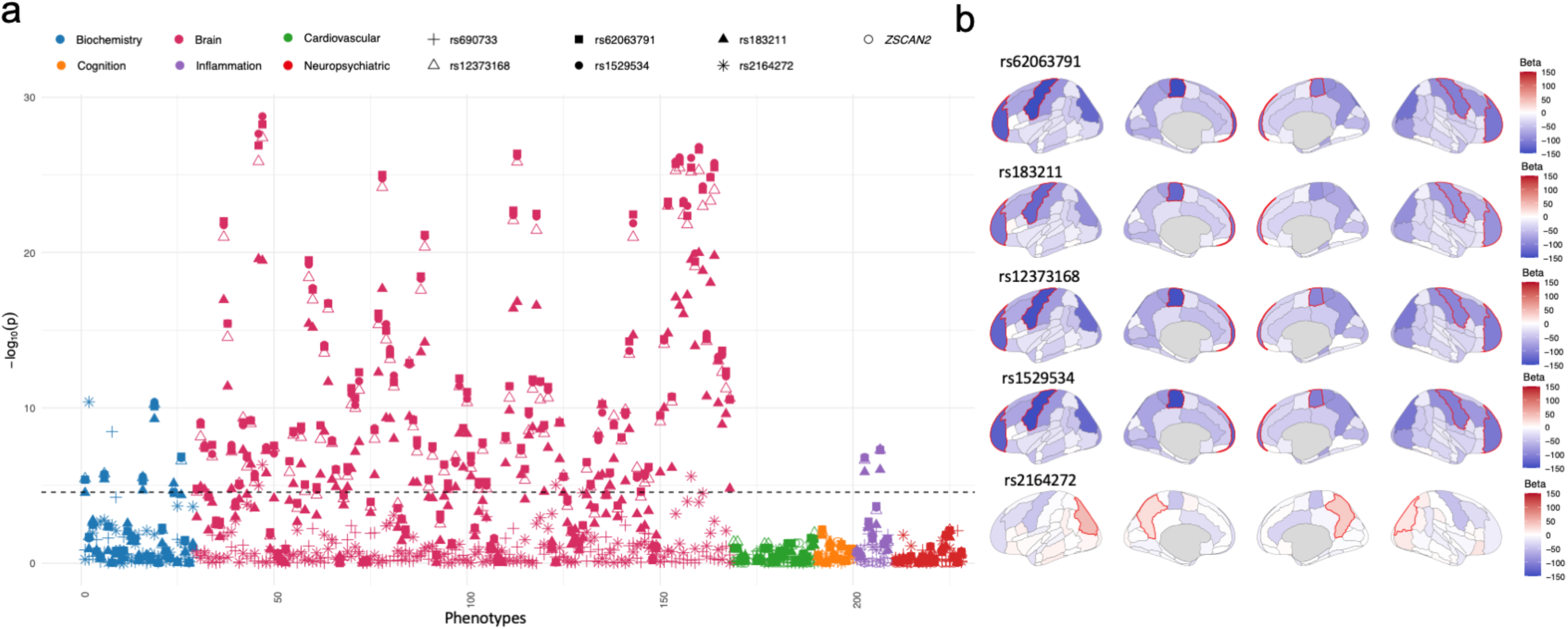
PheWAS and brain region effects of SPARK-supported variants and genes. a, PheWAS results for SPARK-supported variants and genes. Each point represents a mutation type, colored by phenotype category (neurological, brain, cognition, etc.). b, SNPs that remain significant after Bonferroni correction (rs26063791, rs183211, rs12373168, rs1529534, and rs2164722) are shown in brain maps, with beta values for each SNP’s effect on brain region volume (blue: negative, red: positive).

Collectively, these results indicate that autistic traits associated genetic variants preferentially influence brain structure, particularly in cortical regions implicated in social cognition and executive control, providing convergent neurobiological support for their phenotypic effects.

## Discussion

In this study, we present a large-scale exome-wide association analysis of quantitatively measured autistic traits in middle aged and older adults from the general population. By integrating exome sequencing with dimensional behavioral phenotyping in more than 130,000 individuals, we move beyond traditional diagnosis-based frameworks and provide a comprehensive view of the genetic architecture underlying autistic traits across the lifespan especially in the aging population.

A central finding of this study is the strong association signal at the 17q21.31 region for social skill traits. Although multiple genome-wide significant variants were identified, these signals collapse into a single association driven by an extended linkage disequilibrium block, tagged by rs199533. This region is characterized by a well-known inversion polymorphism that generates long-range haplotypes with suppressed recombination. Previous studies have linked this locus to neurodegenerative disorders, cognitive traits, and brain structural variation^18–22^. Here, we extend these findings by demonstrating that variation at this locus is also associated with social skills variation in undiagnosed middle-aged and older adults. Meanwhile, within this locus, several variants were associated with increased scores on the SCQ questionnaire in the SPARK cohort, providing cross-cohort support for their relevance to social behavioral phenotypes. Intriguingly, however, these associations were only replicated in unaffected individuals within SPARK, but not among individuals diagnosed with autism, suggesting that the genetic architecture underlying quantitative variation in autistic traits may be partially distinct from that driving clinical diagnosis. Therefore, our findings highlight the value of moving beyond childhood-centered diagnostic frameworks, demonstrating that population-based analyses of continuously distributed autistic traits in aging adults can uncover genetic signals not captured by conventional disease-focused approaches.

Beyond this locus, two SNPs associated with attention switching in the UK Biobank (rs690733, β = 0.046, P = 4.26 × 10⁻□; rs2164272, β = −0.047, P = 1.73 × 10⁻□) were consistently replicated in SPARK. Notably, the SNP rs2164272 associated with decreased attention switching scores in UK Biobank was also associated with reduced repetitive behavior scores, as measured by the RBSR questionnaire in SPARK. These convergent associations across independent cohorts and phenotypic domains suggest that variation at this locus may influence core dimensions of behavioral flexibility. In particular, rs2164272 maps to the MTMR9 gene, which has been previously implicated in phosphoinositide regulation and endosomal trafficking^29,30^. Given the role of *MTMR9* in phosphoinositide regulation, our findings raise the possibility that *MTMR9* may represent a biologically plausible target for modulating repetitive behaviors.

Beyond that, we identified phenotype-specific associations implicating *SEMA3F* in communication and *EXD1* in attention switching. *SEMA3F* encodes a semaphorin involved in axon guidance and synaptic organization and has previously been implicated in neurodevelopmental processes^23–25^. Our rare-variant analyses further demonstrate that disruptive coding variation contributes substantially to autistic traits in adulthood^5,26,27^. Gene-based analyses identified *ZSCAN2* as key contributors, with evidence that their effects reflect the cumulative impact of multiple rare variants rather than single outlier mutations. The partial age-dependent modulation observed for these associations suggests that rare variants may interact with developmental and aging-related processes, influencing trait trajectories across the lifespan.

The burden heritability analyses revealed a frequency dependent architecture in which ultrarare loss-of-function variants exert the strongest effects, whereas predicted deleterious missense variants contribute more modestly. Moreover, the inverse relationship between rare variant and common variant heritability across AQ domains suggests partially distinct genetic mechanisms underlying different aspects of autistic traits. Social and communicative domains appear to be influenced by both rare disruptive variants and common haplotypic effects, whereas attentional traits are more strongly driven by polygenic background. This dual architecture mirrors observations in clinically diagnosed ASD and supports the continuity between subclinical traits and neurodevelopmental disorders^28^.

By integrating transcriptomic and proteomic data, we further link genetic variation to downstream molecular phenotypes. Tissue and developmental expression analyses highlight preferential enrichment in cortical regions and temporally specific patterns in thalamic, cerebellar, and striatal circuits. In single cell analysis, genes linked to attention switching, such as *MTMR9* and *EXD1*, show weaker enrichment in the mPFC, suggesting that distinct neural substrates may underlie different behavioral dimensions^29^. Protein-wide association analyses demonstrate that replicated variants influence circulating proteins involved in lysosomal function, membrane dynamics, and energy metabolism, providing measurable intermediate phenotypes connecting genetic variation to systemic biology.

Importantly, neuroimaging analyses revealed that variants associated with autistic traits exert measurable effects on brain structure well into adulthood. Variants influencing social skills were associated with reduced volumes in the prefrontal cortex and precentral gyrus, regions critically involved in social cognition, executive control, motor planning, and social communication. Notably, the precentral gyrus corresponds to the primary motor cortex (M1), a region that has recently emerged as a promising therapeutic target in autism. A recent randomized clinical trial demonstrated that non-invasive stimulation of M1 significantly improved social communication deficits in autistic individuals^30^. The observation that social skill associated genetic variants converge on a cortical region that is itself responsive to therapeutic neuromodulation provides an intriguing link between genetic liability, brain structure, and potential intervention strategies. More importantly, these structural effects remain detectable in later life, indicating that the neurobiological consequences of autism related genetic variation persist far beyond early neurodevelopment and continue to shape brain organization throughout ageing.

Our findings support a lifespan model of autism-related behavioral variation. Rather than viewing autism exclusively as a childhood-onset diagnostic category, the present results indicate that genetic influences on social communication, behavioral flexibility, and attentional traits continue to shape individual differences throughout adulthood and ageing. This perspective suggests that autistic traits represent enduring dimensions of human behavioral variation whose biological foundations remain detectable across the lifespan.

However, several limitations should be acknowledged. First, the UK Biobank cohort is not fully representative of the general population. Second, rare variant replication was limited by sample size in external cohorts. Third, our analyses focused on coding variation and did not capture regulatory non-coding elements. Future studies integrating whole genome sequencing, longitudinal phenotyping, and more diverse populations will be essential to refine these findings.

In conclusion, this study establishes a comprehensive genetic framework for autistic traits in later life, demonstrating that subclinical variation in social, communicative, and attentional behaviors is shaped by both shared and distinct genetic mechanisms relative to diagnosed ASD. By shifting the focus from childhood diagnosis to lifelong dimensional traits, our work provides a foundation for understanding autism as a dynamic, lifelong condition and highlights the importance of extending genetic research beyond traditional clinical boundaries.

## Method

### Study population and phenotypes

The UK Biobank is a large prospective cohort study with comprehensive phenotypic and genetic data from approximately 500,000 participants aged 40-69 years at recruitment. The UK Biobank study received ethical approval from the North West Multi-centre Research Ethics Committee (https://www.ukbiobank.ac.uk/learn-more-about-uk-biobank/about-us/ethics), and all participants provided written informed consent. For the current analysis, we used data on demographic characteristics, genetic information, and the AQ questionnaire scores. The AQ is a 50-item self-report measure assessing autistic traits across five domains: social skills (items 1, 11, 13, 15, 22, 36, 44, 45, 47, 48); attention switching (items 2, 4, 10, 16, 25, 32, 34, 37, 43, 46); attention to detail (items 5, 6, 9, 12, 19, 23, 28, 29, 30, 49); communication (items 7, 17, 18, 26, 27, 31, 33, 35, 38, 39); and imagination (items 3, 8, 14, 20, 21, 24, 40, 41, 42, 50). The total AQ score (range 0-50) and subscale scores were derived by summing the corresponding items.

We restricted our analysis to participants aged 40 years or older at baseline. Individuals with a clinical diagnosis of ASD based on hospital records or self-report were excluded to focus on subclinical autistic traits in the general population. To ensure data completeness, we included only participants who provided valid responses to all 50 AQ items; any individual with missing responses was excluded from the analysis.

To validate our findings, we performed replication analyses using data from the SPARK cohort (Simons Powering Autism Research), which includes individuals with clinically confirmed ASD diagnoses. This independent validation cohort allowed us to assess whether genetic variants associated with subclinical autistic traits in the general population also contribute to clinical ASD risk.

### Exome sequencing and data quality control

Whole-exome sequencing was performed on 454,756 participants from the UK Biobank using the IDT xGen Exome Research Panel v.1.0. In addition to central quality control performed by UK Biobank, we implemented rigorous genotype-level, variant-level, and sample-level quality control procedures. Briefly, multi-allelic sites were split into bi-allelic sites for analysis, and calls with low genotype quality or extreme read depth were removed. We excluded monomorphic variants and those failing standard filters, including call rate <90% and deviation from Hardy-Weinberg equilibrium (P ≤ 1×10−15). For sample-level quality control, we removed individuals who had withdrawn consent, were duplicate samples, showed discordance between self-reported and genetically inferred sex, or exhibited anomalous call rates or related metrics.

Following these steps, we applied additional filters to the variants. For single-variant association analysis of common variants, we retained variants with minor allele frequency (MAF) ≥0.01, genotype missingness <2%, and Hardy-Weinberg equilibrium P > 1×10−15. For gene-based burden analysis of rare variants, we retained variants with MAF <0.01 and genotype missingness <2%. After integrating genetic data with phenotypic exclusions (participants aged <40 years, those with ASD diagnoses, and those with incomplete Autism-Spectrum Quotient responses), our final analysis dataset comprised 139,605 unrelated individuals of European ancestry aged ≥40 years with complete phenotypic and high-quality genetic data.

### Variant annotation

We performed variant annotation using ANNOVAR to identify functional consequences^31^. Rare variants (minor allele frequency <1%) were classified based on their predicted functional impact. Loss-of-function (LoF) variants were defined as those annotated as stop-gained, frameshift insertions/deletions or essential splice-site variants. Missense variants were further classified based on the MPC score, a quantitative measure of deleteriousness that integrates evolutionary conservation and structural impact. For gene-based burden analysis, we created four distinct variant sets: (1) LoF variants: All predicted loss-of-function variants, (2) Missense variants with MPC ≥ 2: Missense variants predicted to be deleterious (MPC score ≥ 2), (3) Combined LoF + missense (MPC ≥ 2): Aggregate set including both LoF and deleterious missense variants.

### Effects of genetic variants on autistic traits

To systematically evaluate how different classes of genetic variants influence autistic traits, we conducted burden analyses across multiple variant categories with varying predicted functional impact. Within these genes, we performed burden tests for 4 distinct variant sets: (1) LoF variants, (2) Deleterious missense variants (MPC ≥ 2), (3) Combined LoF + missense (MPC ≥2), (4) Synonymous variants, for each variant set, we calculated the total burden of rare variants per individual and tested for association with the total AQ score and its five subscale scores (social skills, attention switching, attention to detail, communication, and imagination).

### Exome-wide association analysis

To comprehensively assess genetic associations with autistic traits, we performed both single-variant and gene-based analyses using REGENIE. For single-variant association tests, we included all variants with minor allele count ≥20 and applied inverse normalization to quantitative traits (total AQ score and subscale scores). All models were adjusted for age, sex, and the top 10 principal components of genetic ancestry. Variants with P < 5×10^-8^ were considered genome-wide significant. For gene-based burden analysis, we implemented SKAT-O tests within REGENIE to evaluate the aggregate effects of rare variants (MAF <1%). We applied two MAF thresholds (1% and 0.1%) and tested four variant annotation groups: LoF, missense (MPC ≥ 2), LoF + missense (MPC ≥ 2), Synonymous. Gene-based associations were considered statistically significant after Bonferroni correction for multiple testing. Additionally, we tested for interactions between genetic variants and age or sex by including multiplicative interaction terms in the regression models. This allowed us to assess whether genetic effects on autistic traits varied across age groups or between males and females.

### MAGMA analysis

MAGMA analyses were conducted using MAGMA implemented in the FUMA platform (https://fuma.ctglab.nl/). Summary statistics from the exome-wide association analyses were uploaded to FUMA and analyzed using default parameters^32^.

### Validation of UKB-identified common and rare variants in the SPARK cohort

To assess the robustness and generalizability of genetic associations identified in the UK Biobank, we performed independent validation analyses in the SPARK autism cohort using SPARK-specific phenotypic measures. A total of 65 common Snps identified from autistic traits analyses were selected for validation. Given differences in phenotypic instruments between cohorts, a domain-matched validation strategy was adopted. Specifically, Snps associated with AQ social skill and communication subdomains were evaluated using the Social Communication Questionnaire total score, whereas Snps associated with AQ attention-related traits were evaluated using the Repetitive Behavior Scale–Revised total score.

For each SNP, individuals in SPARK were classified as carriers or non-carriers of the effect allele. Mean differences in phenotypic scores between groups were estimated, and corresponding effect sizes and confidence intervals were calculated. Statistical significance was assessed using two-sample tests without assuming equal variances.

Rare LoF variants in *ZSCAN2* and *NBR1*, identified in UK Biobank rare variant burden analyses, were evaluated in SPARK. Due to the small number of LoF carriers for each gene, standard parametric testing was considered underpowered. Therefore, we applied a permutation-based testing framework, in which carrier labels were randomly permuted while preserving group sizes to generate empirical null distributions of mean phenotype differences. Empirical P values were calculated as the proportion of permuted statistics exceeding the observed mean difference. Cohen’s d was reported to facilitate effect size interpretation.

### LOVO analysis

To assess the robustness of significant gene-based associations, we performed LOVO analysis. For each significant gene, we iteratively removed one variant at a time and recalculated the association P value. Variants whose removal substantially altered the association signal (ΔP > 1 order of magnitude) were identified as potential drivers of the gene-level association. This approach helped distinguish genes where multiple variants contribute to the signal from those driven by a single influential variant.

### Conditional analysis of common variant signals

To determine whether the observed association signals were driven by independent common variants within each locus, we performed stepwise conditional analyses. For each locus of interest, we defined a genomic region spanning ±500 kb around the lead signal. Within this region, we first conducted single-variant association analyses of all common variants (MAF >1%) using PLINK2, adjusting for age, sex, and genetic principal components. We then applied LD clumping (P₁ < 5×10⁻□, P₂ < 1×10⁻□, r² < 0.1, window size = 2 Mb) to identify approximately independent lead SNPs. Subsequently, we performed stepwise conditional analyses using REGENIE (step 2), iteratively including the genotype dosage of each identified lead SNP as covariates in the model. Conditional analyses were repeated until no additional genome-wide significant signals remained. Variants that remained significant after conditioning on previously identified lead SNPs were considered independent association signals.

### Conditional analysis for rare variant signals

To determine whether the identified rare variant associations were independent of nearby common genetic variation, we performed conditional analyses. For each gene that showed significant association in the gene-based burden test (P < 2.5×10⁻□), we defined a genomic region spanning 500 kb upstream and downstream of the gene. Within this region, we first performed single-variant association analysis for all common variants (minor allele frequency >0.5%) with the target autistic trait using Plink2. We then applied linkage disequilibrium clumping (clumping P threshold <1×10⁻□, r² threshold <0.01) to identify independent common variant signals. Finally, we re-ran the gene-based burden test for the target gene, adjusting for the genotypes of these clumped, independent common variants as covariates in the model. A rare variant association that remained significant after conditioning on nearby common variants (P < 2.5×10⁻□) was considered an independent genetic signal.

### Heritability and genetic correlation of rare variant burden

To quantify the aggregate contribution of rare coding variants to autistic traits and assess the genetic sharing across traits, we estimated the heritability explained by gene-wise burden scores (h²_burden) and their genetic correlations using the BHR tool. Variants were stratified by allele frequency into ultra⍰rare (MAF < 1×10⁻□) and rare (1×10⁻□ ≤ MAF < 1×10⁻³) bins, and by functional consequence (LoF and missense with MPC ≥ 2). Summary statistics for burden association tests generated by REGENIE were used as input.

First, univariate BHR analysis was performed to estimate h²burden for each autistic trait (total AQ score and five subscale scores). We aggregated the heritability estimates from ultra⍰rare and rare bins to obtain the total burden heritability for each variant category. These estimates were then compared with common⍰variant heritability (h²SNP) for the same traits derived from linkage disequilibrium score regression using publicly available GWAS summary statistics. Second, bivariate BHR analysis was conducted to estimate genetic correlations (rg): Within⍰trait, across variant categories: between LoF burden and missense burden for the same trait. Across traits, within variant categories: between different autistic trait scores for LoF and missense burdens separately.

### Gene-set enrichment and tissue expression analysis

To characterize the biological functions and tissue expression patterns of the identified genes, we performed functional enrichment analyses using the clusterProfiler R package. The analyzed gene set comprised 28 prioritized genes, including genes harboring SPARK-replicated common variants, genes identified from SPARK-validated rare-variant analyses, and genes prioritized by MAGMA. GO enrichment analyses were conducted across three categories: Biological Process, Cellular Component, and Molecular Function. In parallel, tissue-specific expression enrichment was evaluated using transcriptomic data from the GTEx project to assess whether the prioritized genes showed preferential expression in tissues relevant to brain function and neurodevelopment^33^. To further investigate developmental and regional expression patterns, we analyzed transcriptomic data from the BrainSpan atlas^34^. Genes associated with the social skill and communication domains—two core dimensions of autistic traits—were examined across multiple developmental stages and brain regions, allowing assessment of spatiotemporal expression dynamics during human brain development.

### Single-cell expression analysis

To identify neural cell types in which autistic trait–associated genes are preferentially expressed, we performed single-cell transcriptomic analyses using publicly available human brain data. We obtained single-nucleus RNA sequencing data from the GEO under accession number GSE97930. Only adult samples derived from the frontal cortex (Brodmann area 10, BA10), corresponding to the medial prefrontal cortex (mPFC), were included in downstream analyses.

Raw UMI count matrices (GSE97930_FrontalCortex_snDrop-seq_UMI_Count_Matrix_08-01-2017.txt) were used as input for all analyses. Data preprocessing, quality control, normalization, and downstream analyses were performed using the Seurat R package (v5.0.1)^33^. Differentially expressed marker genes for each cluster were identified using Seurat’s FindAllMarkers function. For each cluster, the top 50 marker genes ranked by average log2 fold change were selected for cell-type annotation.

### Phenome-wide association analysis

To characterize the broader phenotypic impact of the identified genes, we performed PheWAS analysis across a comprehensive range of diseases, cognitive measures, and physiological traits available in the UK Biobank^35^ (Table S2). The analysis encompassed six major domains: (1) neuropsychiatric disorders; (2) cardiovascular diseases; (3) cognitive performance; (4) brain MRI-derived phenotypes; (5) biochemical markers; and (6) inflammatory markers. Only high-confidence variants and genes that were independently validated in the SPARK cohort, including common-variant SNPs and rare-variant gene-level associations, were included in the analyses. This approach enabled systematic assessment of the pleiotropic effects of autistic trait–associated loci and their potential contributions to comorbid conditions and systemic physiology.

## Supplemental Information

Supplementary Figures.pdf Figures S1-S18

Supplementary Tables.xlsx

Tables ST1-ST12

## Data availability

The individual-level phenotypic and genetic data used in this study were accessed from the UK Biobank (https://www.ukbiobank.ac.uk/) under application number 217390. The genomic and phenotypic data for the SPARK (https://www.sfari.org/resource/spark/) are available at SFARI Base (iWESv3, https://base.sfari.org/). Single-cell RNA sequencing data from the human brain were obtained from the Gene Expression Omnibus under accession number GSE97930, and analyses were restricted to cells from the mPFC.

## Acknowledgments

This work was supported by the Brain Science and Brain-like Intelligence Technology-National Science and Technology Major Project (2025ZD0218000), the National Natural Science Foundation of China (82471194), and the “Fundamental Research Funds for the Central Universities” starting fund (BMU2022RCZX038) to T.W.

## Declaration of Interests

The authors declare no competing interests.

---

**Table 1.** Exome-wide significant SNPs associated with autistic traits and validated in the SPARK cohort (*P* < 5 × 10^-8^).

| Traits | Chr | rsID | A0 | A1 | A1freq | Beta | Se | P-value | Gene | SPARK | SPARK_Scale | SPARK_Beta | MAGMA |
| --- | --- | --- | --- | --- | --- | --- | --- | --- | --- | --- | --- | --- | --- |
| social_skill | 17 | rs183211 | G | A | 0.2403 | 0.0685 | 0.0116 | 3.21E-09 | NSF | Yes | SCQ | 0.3414 | Yes |
| social_skill | 17 | rs1529534 | A | G | 0.2214 | 0.0739 | 0.0119 | 5.31E-10 | MAPT | Yes | SCQ | 0.4879 | Yes |
| social_skill | 17 | rs62063791 | A | G | 0.2214 | 0.0733 | 0.0119 | 7.16E-10 | MAPT | Yes | SCQ | 0.4780 | Yes |
| social_skill | 17 | rs12373168 | A | C | 0.2243 | 0.0737 | 0.0118 | 4.86E-10 | MAPT-AS1 | Yes | SCQ | 0.2360 | No |
| attension_swiching | 8 | rs2164272 | A | C | 0.4077 | -0.0471 | 0.0083 | 1.73E-08 | MTMR9 | Yes | RBSR | -0.6269 | Yes |
| attension_swiching | 15 | rs690733 | G | A | 0.4314 | 0.0455 | 0.0083 | 4.26E-08 | EXD1 | Yes | RBSR | 1.0376 | No |

**Table 2.** Rare-variant gene-based association results for autistic traits.

| Traits | Chr | Gene | Group | Max MAF | Beta | Se | SKAT-O | SPARK |
| --- | --- | --- | --- | --- | --- | --- | --- | --- |
| AQ_total | 17 | NBR1 | LoF | 0.00111827 | 1.76056 | 0.386046 | 5.10E-06 | No |
| communication | 15 | ZSCAN2 | LoF | 0.000175564 | 1.00278 | 0.272555 | 0.000234 | No |

## Notes

### Competing Interest Statement

The authors have declared no competing interest.

### Author Declarations

The study was reviewed with ethical approval given by the Biomedical Ethics Committee of Peking University (No. PUIRB-YS2023128).

